# Surface Electromyographic Assessment of Pelvic Floor Muscles Protocol Tested by a Novel Airbag-type Stretchable Electrode Array (ASEA) Device in Stress Urinary Incontinence in Postmenopausal Women

**DOI:** 10.1101/2025.01.26.25321148

**Authors:** Zujuan Zhang, Qian Chen, Suyun He, Wenjuan Li, Shengming Wang, Zhenwei Xie

## Abstract

Electromyography (EMG) is a well-established method to quantify the PFM activity. The purpose of this study was to investigate the PFM of US and LAMs EMG variables separately by ASEA device and determine the predict value of sEMG for ASEA assessing the PFM in postmenopausal women with SUI. We obtained the EMG of US and LAMs separately by ASEA device consisted of following elements: ARP, MVC, TCP, ECP and PRP. We performed ROC curve analysis for optimal cutoff level for sEMG activation of the PFM. For 65 healthy and 67 with SUI, muscle strength was lower in SUI group according to the modified Oxford grading system by bidigital palpation. The sEMG of PFM activities were significantly lower in SUI group tested by ASEA device, especially in the MVC, TCP and ECP activities in US and LAMs. According to ROC curves, US and LAMs, especially PR and PC, except IC, have optimum predict cutoff amplitude for SUI. Our study proved that US defect and dysfunctional LAMs play an important role in the pathogenesis of SUI and separation of specific regions of US and LAMs can provide a reliable and optimized treatment strategy for the precise rehabilitation of pelvic floor muscles.

## 1. Introduction

SUI is defined as involuntary loss of urine on effort or physical exertion[1]. Stress urinary incontinence (SUI) refers to the involuntary leakage of urine from the external urethral orifice when abdominal pressure increases due to actions such as sneezing, coughing and laughing.The prevalence of SUI has been reported to be as high as 49%, depending on population and definition[2]. The incidence of SUI in postmenopausal women is even worse, which is as high as 57%-75%[3]. Approximately 25% of young women, 44–57% of middle-aged and postmenopausal women, and 75% of older women experience some involuntary urine loss[4].It seriously affects the physical and mental health of the patients and also represents a significant medical and economic burden on society[5-7].

Currently, supervised pelvic floor muscle training (PFMT) of at least 3months ‘ duration is the first-line treatment for SUI[3,6].PFMT involves the regular practice of repeated voluntary pelvic floor muscle contractions, with sufficient exercise progression, in order to produce a training effect on the muscles. Surface electromyography (sEMG) is considered an useful tool for real-time evaluation of pelvic floor muscle (PFM) contractions and for the assessment of PFM function by identification of the PFM motor unit action potential[3]. The validity of the use of the surface EMG (sEMG) in the assessment of bioelectrical activity of PFM as well as muscles that act as synergistic muscles to PFM has been proven by many studies[8-10].Electrical signals from the muscles are generated by the recruitment of motor units during contraction; a correlation has been observed between muscle strength and the activation of motor units.

While the electrical information of pelvic floor muscles are measured by an internal vaginal probe with a hard two-channel electrode, which is universally used in clinic work. Such two-channel electrode procedure is simple, fast and easy to operate. However, Such surface EMG does not determine precisely which muscles are being measured, but rather sum the activity from all muscles coming into contact with the electrode plates. Furthermore with the characters of unsuitable bonding interface strategy, traditional two channel electrode distribution and lack of proper and accurate evaluation method, it is difficult to obtain accurate state of the muscles under specific conditions. We propose a novel airbag-type stretchable/inflatable electrode array (ASEA) device for sEMG signal acquisition for PFM analysis. As we previously reported, the ASEA can inflate to match the variously sized vaginal cavity of different human bodies, while following the muscle movement, which provides a stable probe-muscle contact interface and has great biological adaptability[11]. Twenty-four differential pair sEMG collection channels form 10 typical PFM cover regions according to the approximate muscle fiber extension direction. The covered PFM regions include urethral sphincter (US), vaginal sphincter (VS), external anal sphincter (EAS), Levator ani muscle (LAM), which include puborectalis (PR), pubococcygeus (PC) and iliococcygeus (IC) muscles.

Such electrode distributed along the muscle fiber ensures the accuracy of signal acquisition, and the regionalization division makes it possible to pinpoint the specific muscle regions.

Benefit from ASEA’s stable and accurate sEMG signal acquisition, several key parameters are proposed to comprehensively evaluate regional muscles which has close relationship of SUI, the evaluations include: pre-baseline, it means anterior resting potential(ARP), maximum voluntary contraction(MVC),it means rapid contraction potential(RCP), tonic contraction potential(TCP), endurance contraction potential(ECP) and post-baseline, it means posterior resting potential(PRP) was calculated according to the regional function results.The aim of the current study was to evaluate the characteristics of specific and precise position of PFM sEMG during rest and contraction by ASEA device methods in postmenopausal women with SUI and in healthy women.

## 2. Materials and methods

### 2.1 Ethics statement

Ethical approval for the present study was granted by the Ethics Committee of the School of Medicine of Zhejiang University,Hangzhou, China(No. IRB-20220008-R &No. IRB-20240381-R). All participants provided written informed consent to participate in the study, and the ethics committee approved the consent procedure.

### 2.2 Study design

The current study was designed as a retrospective data analysis, approved by the Ethics Committee of Women’s Hospital School of Medicine, Zhejiang University (China, No.IRB-20220008-R). The data for the current study were originally sampled by the same research group focusing on the effect of whole-body vibration on PFM activity.

### 2.3 Participants

For the purpose of the current study, 2 groups were determined: 65 volunteers healthy postmenopausal women, such postmenopausal women had no history of pelvic floor dysfunction. 67 postmenopausal women with stress urine incontinence (SUI) from Women’s Hospital School of Medicine, Zhejiang University. The study was conducted from December 2021 to December 2024.Women were recruited from January 2022 to September 2023 (Fig 1 & Table 1). The postmenopausal women with SUI should include such inclusion criteria:(1) Clinical diagnosis of SUI;(2)Have a history of sex; (3)Menopause≥1 year. While the exclusion criteria were as followings : (1)Acute reproductive organ inflammation;(2)With cardiac pacemakers; (3)Malignant tumors; (4)History of pelvic radiotherapy; (5)Pelvic floor surgery≤6 months; (6)Any disease or symptom that may affect the implementation of the study or the interpretation of the results; (7)Participate in other clinical trials at the same time.

**Table 1.** Demographics of participants.

| Characteristics | Group |  | t | Z | P |
| --- | --- | --- | --- | --- | --- |
|  | SUI<br>n=67 | Healthy<br>n=65 |  |  |  |
| Age (years) | 58.73±6.43 | 57.38±6.01 | 1.24 | / | 0.216 |
| BMI | 23.68±3.16 | 23.01±2.46 | 1.37 | / | 0.175 |
| WHR (waist-hip ratio) | 0.91±0.60 | 0.89±0.06 | 1.85 | / | 0.067 |
| Years after menopause (years) | 7(4, 10) | 6(3,10.5) | / | 0.79 | 0.428 |
| Parity | 3(2, 3) | 2(2, 4) | / | 1.31 | 0.189 |
| Gravidity | 1 (1, 2) | 1 (1, 2) | / | 0.60 | 0.546 |
| Vaginal delivery | 1 (1, 2) | 1 (1, 2) | / | 0.15 | 0.878 |
| Cesarean section | 0 (0, 0) | 0 (0, 0) | / | 1.14 | 0.253 |

*Data are mean ± SD unless indicated. Healthy: women with healthy pelvic floor muscles; SUI: women with stress urinary incontinence. $p < 0.05$ means have statistical significance. T : Statistical value of the T-test; Z: Statistical value of Wilcoxon test.*

**Figure 1.**
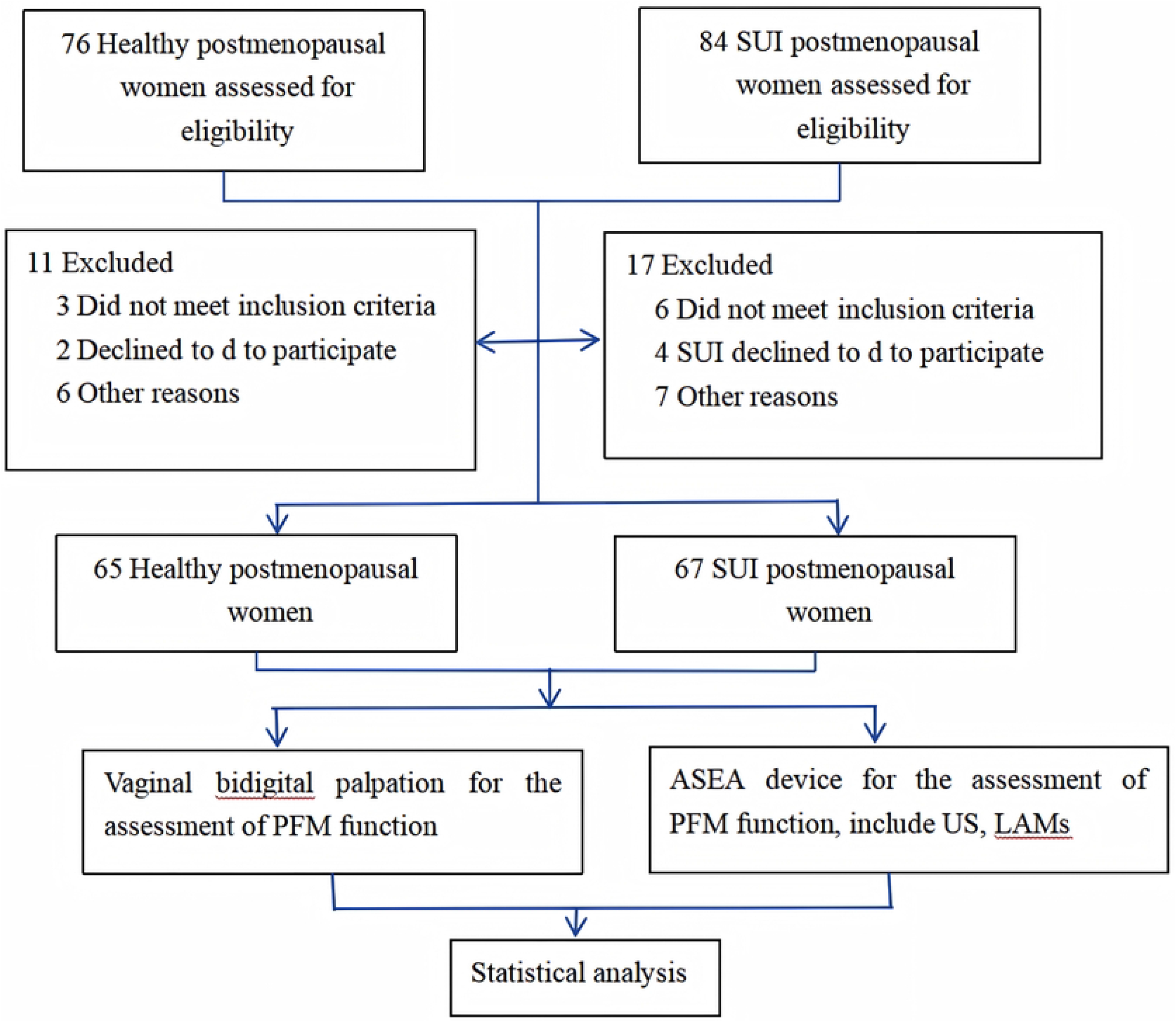
Consolidated Standards of Reporting Trials (CONSORT) Diagram

### 2.4 Procedures and data collection

#### 2.4.1 Vaginal bidigital palpation for the assessment of PFM function

PFM function assessment by vaginal bidigital palpation (PERFECT scheme)[12,13]during which the participants were taught to properly contract their PFM and asked to perform a maximal PFM contraction.

All the postmenopausal women tested a PFM testing score of M0–M5 according to the modified Oxford grading system:

M0: no contraction;

M1: flicker;

M2: weak;

M3: moderate;

M4: good;

M5: strong

#### 2.4.2. ASEA device for the assessment of PFM function

##### 2.4.2.1. Introduction of airbag-type stretchable electrode array (ASEA) device

The ASEA is an airbag-type inflatable probe which is able to provide a tight contact and stable interface between electrode units and PFM. As we had published previously, ASEA has designed 24 bipolar electrode pairs (totally 32 contact pads) placed along the muscle fiber directions, then forming 10 typical PFM regions, thereby allowing to collect high-quality sEMG signals[11].The local flexible circuit image under microscope and explosive structure of ASEA is shown in Figure 2a, and the physical object of ASEA is shown in Figure 2b. the ASEA device can obtain the electrical potential distribution and the corresponding frequency information for all the major muscles in vagina, which include: urethral sphincter (US), vaginal sphincter (VS), external anal sphincter (EAS), Levator ani muscle (LAM), which is composed of puborectalis (PR), pubococcygeus (PC) and iliococcygeus (IC) muscles. The details of fabrication process, arrangement of the electrodes (electrical contacts), clinical reliability evaluation of ASEA device can refer to our previous paper[14], and the results have demonstrated that the accuracy and stability of ASEA device are far superior to existing PFM electrode probes.

**Figure 2.**
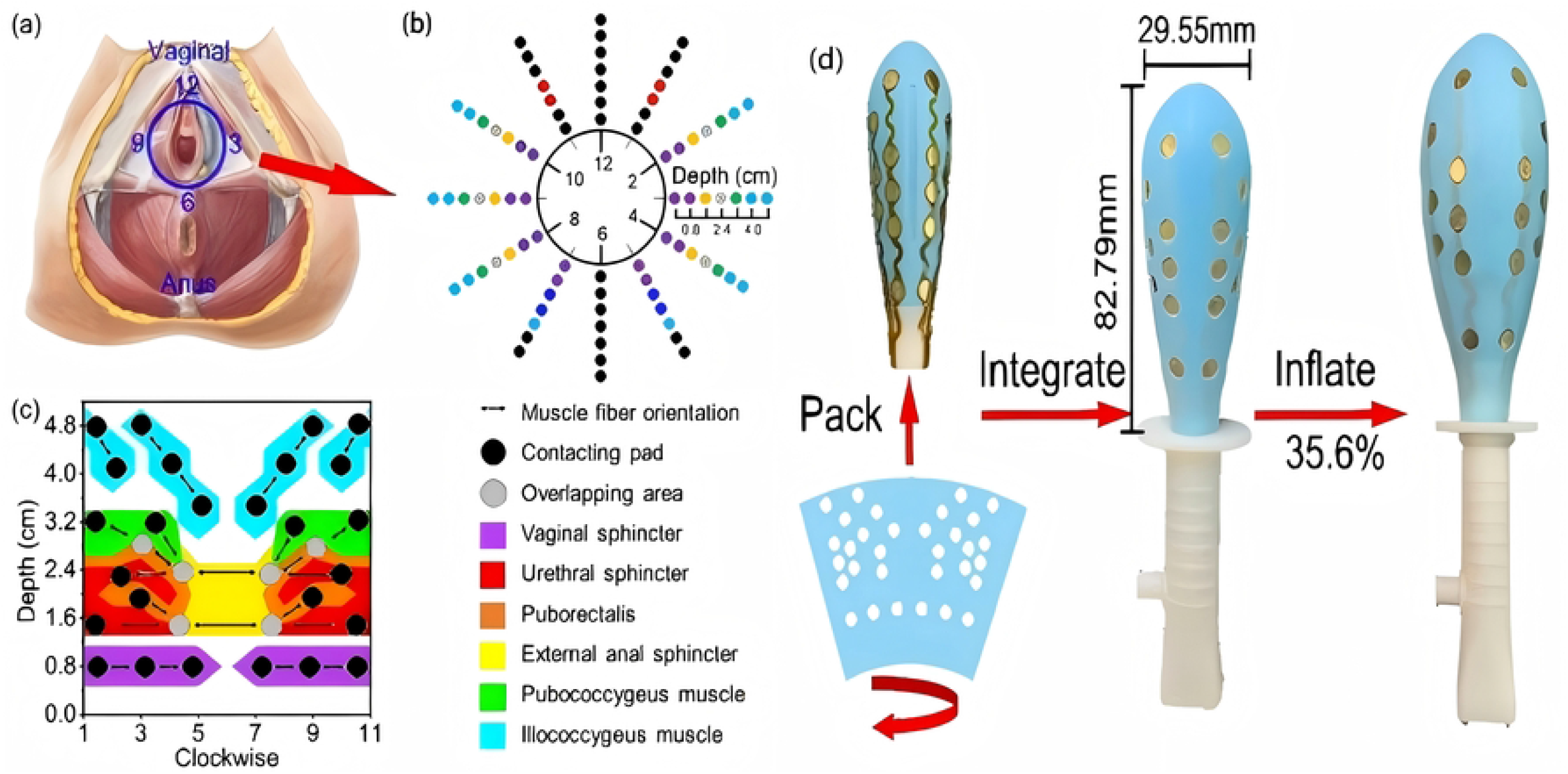
Structure and configuration of the ASEA probe. (a). The distributions of pelvic muscles. (b).The details of the double-layer electrode unit. Macro internal structure of ASEA (left), the ASEA probe before inflation (middle) and after inflation (right).(c). Distribution of main pelvic muscles in a 2D format and locations of the contact pads of ASEA (black dots) that are distributed according to the major muscle distribution and muscle fiber directions and unit (arrow). Each muscle is marked with specific color for better distinguishment. (d) The shape and formation of ASEA.

##### 2.4.2.2 Processure of ASEA device for the assessment of PFM function

Women were first instructed in the correct PFM activation in supine position with slightly bent knees and neutral hip position, which was controlled with electromyographic biofeedback by a therapist experienced in pelvic floor rehabilitation. The same therapist performed all assessments to standardize the procedures. PFM activity was measured with a novel airbag-type stretchable electrode array (ASEA) probe (Patent No. ZL 2022 1 0927100.3,China). Patients were asked first to squeeze their PFMs freely to get used to the test probe. Abdomen, perineum, hip and gluteus muscles were simultaneously observed to ensure an isolated activation of PFM. Afterwards,the ASEA probe was carefully inserted into the vagina, with the placement controlled before measurement. The protocol of all measurements of PFM activity consisted of the assessment of the following elements: “pre-baseline” (10 s PFM activity at rest before functional measurements), “flick contractions” was also means maximum voluntary contraction(MVC) (10 s measurement, in which participants performed short, quick contractions of the PFM), “tonic contractions” (5 × 10 s contractions, in which the participant tried to contract the PFM and hold for 10 s), “endurance contraction” (in which the participant attempted to hold the PFM contraction for 60 s), and “post-baseline” (10 s PFM at rest after functional measurements).

### 2.5 Statistical analysis

Statistical analysis was conducted using SPSS 26.0. The distributions of baseline characteristics in the SUI group and the non-SUI group were compared with the chi-squared test for proportions and either the t-test for continuous variables. Univariable and multivariable logistic regression were used to evaluate the influence of individual predictor variables, as well as their combined effect in predicting the absence of SUI. Statistically significant predictors were subjected to a backward, stepwise logistic regression analysis to determine which combination of predictors best explained the response. To determine the optimal cutoff level for sEMG activation of the PFM to detect the occurrence of SUI, we performed receiver operating characteristic (ROC) curve analysis.. For all comparisons, we defined α as 0.05.

## 3. Results

### 3.1 The Demographic and Clinical Characteristics

In total, 132 post menopausal women were recruited and screened for this study. And 67 in the SUI group, 65 in the healthy group. The demographic and clinical characteristics were as the followings(Table 1). There was no statistical significance in the two groups of post menopausal women in age, BMI, WHR, gestation, and mode of deliver(*p > 0*.*05*).

### 3.2 PFM function assessment by vaginal bidigital palpation

All the postmenopausal women in the two group firstly tested a PFM testing score of power M0– M5 according to the modified Oxford grading system,the results were as followings(Table 2). The score of power of SUI group in post menopausal women was lower than healthy women(*p < 0*.*05*).

**Table 2.** PFM function assessment by vaginal bidigital palpation.

| muscle strength | Group |  | Z | P |
| --- | --- | --- | --- | --- |
|  | SUI<br>n=67 | Healthy<br>n=65 |  |  |
| type-I muscle fiber (power 0-5) | 1 (1, 2) | 2 (1, 3) | 4.48 | < 0.001 |
| type-II muscle fiber | 1 (1, 2) | 2 (1.5, 3) | 4.53 | < 0.001 |

*Healthy: women with healthy pelvic floor muscles; SUI: women with stress urinary incontinence. $p < 0.05$ means have statistical significance*

### 3.3 Comparison of surface electromyography (sEMG) results between SUI and non-SUI women

Multi-channel sEMG signals were successfully acquired from all participants.The relationships between the absence of SUI and the sEMG activity of the PFM were determined by t-test for continuous variables. (Table 3). The PFM activity included as followings:”ARP (anterior resting potential))” (10 s PFM activity at rest before functional measurements), “MVC (maximum voluntary contraction)” (10 s measurement, in which participants performed short, quick contractions of the PFM), “TCP (tonic contraction potential)” (5 × 10 s contractions, in which the participant tried to contract the PFM and hold for 10 s), “ECP (endurance contraction potential)” (in which the participant attempted to hold the PFM contraction for 60 s), and “PRP (posterior resting potential)” (10 s PFM at rest after functional measurements). In the rest station, both in the pre-baseline and post-baseline almost all the PFM showed stable conditions, there is no statistic difference between postmenopausal SUI group and non-SUI group. There is a statistically significant decrease in urethral sphincter (US), puborectalis (PR) and pubococcygeus (PC) during flick contractions, tonic contractions, endurance contraction was noted in the postmenopausal SUI group compared with postmenopausal non-SUI group(Fig 3).

**Table 3.**
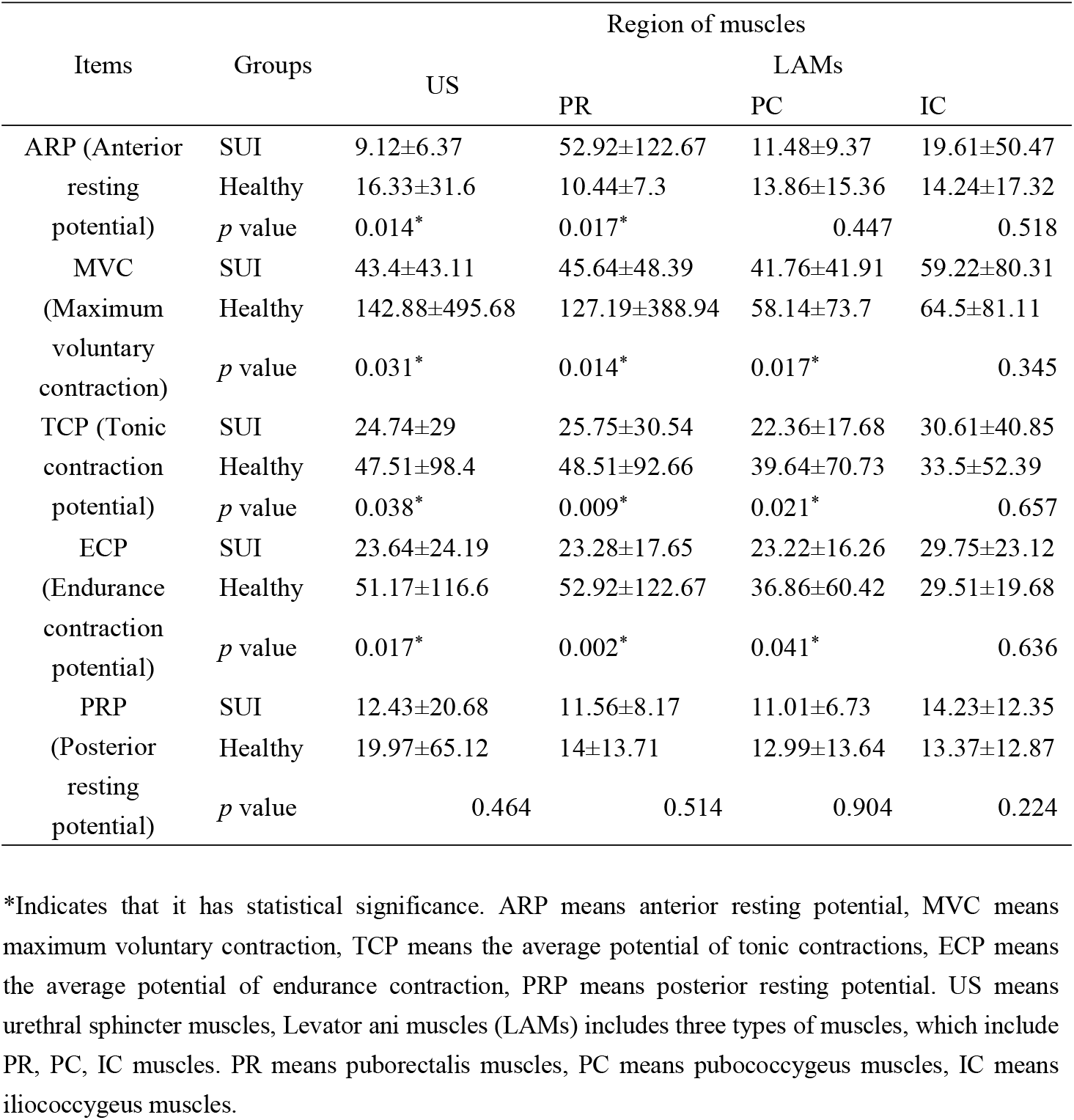
The surface electromyography (sEMG) results of US and LAMs (including PR, PC, IC muscles) in postmenopausal women.

| Items | Groups | Region of muscles |  |  |  |
| --- | --- | --- | --- | --- | --- |
|  |  | US | PR | PC | IC |
| ARP (Anterior resting potential) | SUI | 9.12±6.37 | 52.92±122.67 | 11.48±9.37 | 19.61±50.47 |
|  | Healthy | 16.33±31.6 | 10.44±7.3 | 13.86±15.36 | 14.24±17.32 |
|  | *p* value | 0.014* | 0.017* | 0.447 | 0.518 |
| MVC (Maximum voluntary contraction) | SUI | 43.4±43.11 | 45.64±48.39 | 41.76±41.91 | 59.22±80.31 |
|  | Healthy | 142.88±495.68 | 127.19±388.94 | 58.14±73.7 | 64.5±81.11 |
|  | *p* value | 0.031* | 0.014* | 0.017* | 0.345 |
| TCP (Tonic contraction potential) | SUI | 24.74±29 | 25.75±30.54 | 22.36±17.68 | 30.61±40.85 |
|  | Healthy | 47.51±98.4 | 48.51±92.66 | 39.64±70.73 | 33.5±52.39 |
|  | *p* value | 0.038* | 0.009* | 0.021* | 0.657 |
| ECP (Endurance contraction potential) | SUI | 23.64±24.19 | 23.28±17.65 | 23.22±16.26 | 29.75±23.12 |
|  | Healthy | 51.17±116.6 | 52.92±122.67 | 36.86±60.42 | 29.51±19.68 |
|  | *p* value | 0.017* | 0.002* | 0.041* | 0.636 |
| PRP (Posterior resting potential) | SUI | 12.43±20.68 | 11.56±8.17 | 11.01±6.73 | 14.23±12.35 |
|  | Healthy | 19.97±65.12 | 14±13.71 | 12.99±13.64 | 13.37±12.87 |
|  | *p* value | 0.464 | 0.514 | 0.904 | 0.224 |

\*Indicates that it has statistical significance. ARP means anterior resting potential, MVC means maximum voluntary contraction, TCP means the average potential of tonic contractions, ECP means the average potential of endurance contraction, PRP means posterior resting potential. US means urethral sphincter muscles, Levator ani muscles (LAMs) includes three types of muscles, which include PR, PC, IC muscles. PR means puborectalis muscles, PC means pubococcygeus muscles, IC means iliococcygeus muscles.

**Figure 3.**
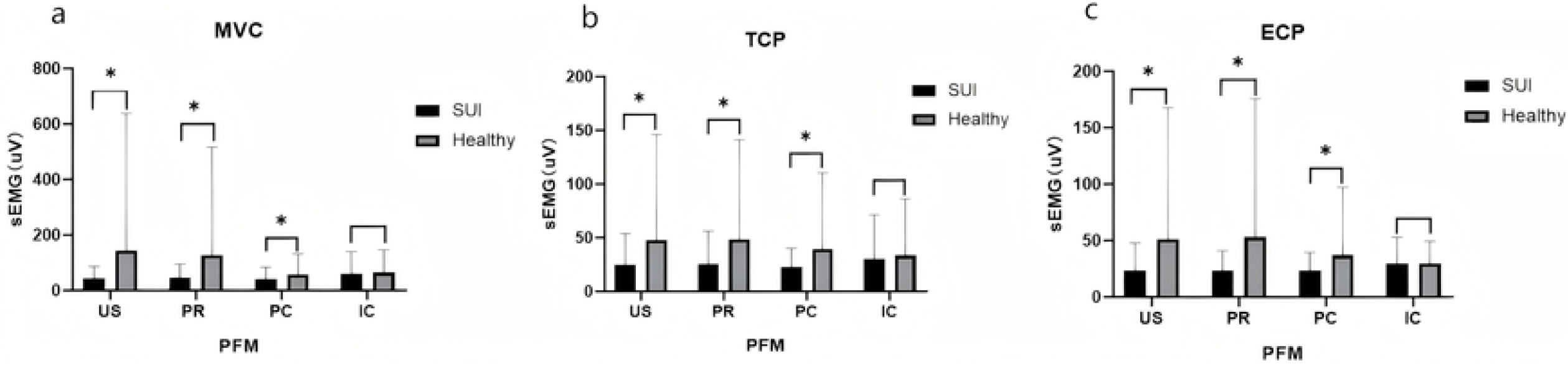
(a) The comparison of MVC in US, PR, PC and IC. During flick contractions (MVC) conditions, there is a statistically significant decrease of amplitude in urethral sphincter (US), puborectalis (PR) and pubococcygeus (PC) in postmenopausal SUI group. (b) The comparison of TCP in US, PR, PC and IC. In tonic contractions, there is a statistically significant decrease of amplitude in urethral sphincter (US), puborectalis (PR) and pubococcygeus (PC) in postmenopausal SUI group. (c) The comparison of ECP in US, PR, PC and IC. In endurance contraction, there is a statistically significant decrease of amplitude in urethral sphincter (US), puborectalis (PR) and pubococcygeus (PC) in postmenopausal SUI group. *Indicates that it has statistical significance.

### 3.4 Prediction Capability of the sEMG

ROC curves were constructed separately for all sEMG tests (Figure 4) in order to present the prediction ability of these variables to predict SUI. The optimum predict cutoff for “Flick contractions (MVC)” of urethral sphincter (US) was 48.55 (area under the curve (AUC), 0.61; 95% confidence interval (CI), 0.51-0.71; sensitivity at 76.1%, specificity 53.8%), “Tonic contractions” of urethral sphincter (US) was 25.79 (AUC, 0.61; 95% CI, 0.51-0.70, sensitivity at 77.6%, specificity 44.6%), “Endurance contraction”of urethral sphincter (US) was 23.02(AUC:0.61,95%CI:0.53-0.72;sensitivity at 70.1%,specificity 60%).

**Figure 4.**
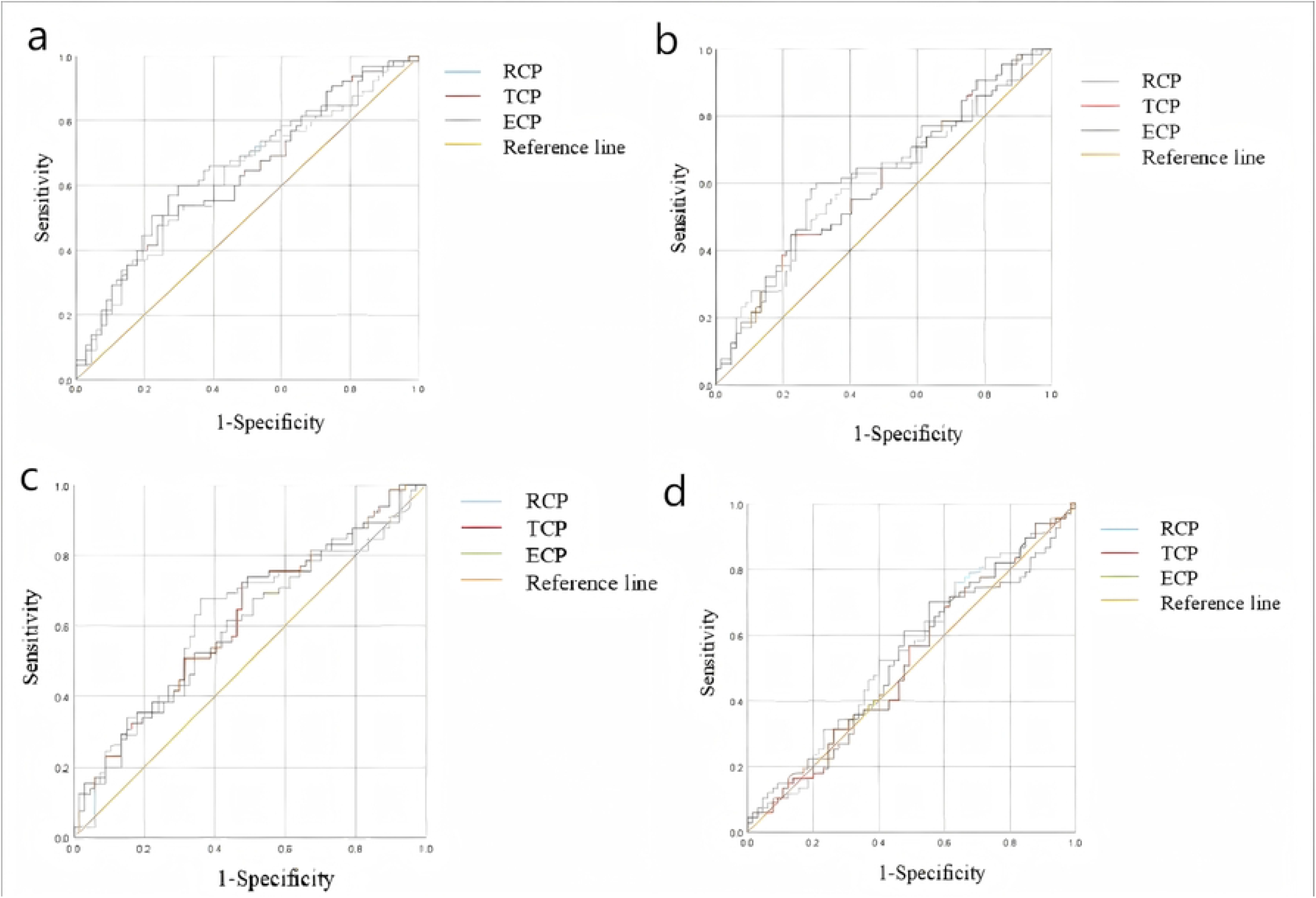
Receiver operating characteristic (ROC) curve (with cutoff point marked) for sEMG activity in the pelvic floor muscle of patients with SUI compared with non-SUI individuals.(a)ROC curves of EMG tests of US.(b)ROC curves of EMG tests of PR.(c)ROC curves of EMG tests of PC. (d)ROC curves of EMG tests of IC.

The optimum predict cutoff for “Flick contractions (MVC)” of puborectalis (PR) was 34.48 (area under the curve (AUC), 0.62; 95% confidence interval (CI), 0.53-0.72; sensitivity at 55.2%, specificity 67.7%), “Tonic contractions” of puborectalis (PR) was 25.96 (AUC,0.63; 95%CI, 0.54-0.73, sensitivity at 74.6%, specificity 50.8%), “Endurance contraction”of puborectalis (PR) was 26.20(AUC:0.65,95%CI:0.56-0.75;sensitivity at 70.1%,specificity 60%).

Pubococcygeus (PC)

The optimum predict cutoff for “Flick contractions (MVC)” of pubococcygeus (PC) was 35.05 (area under the curve (AUC), 0.62; 95% confidence interval (CI), 0.52-0.72; sensitivity at64.2%, specificity 67.7%), “Tonic contractions” of pubococcygeus (PC) was 17.36 (AUC,0.62;95%CI, 0.52-0.71, sensitivity at 50.7%, specificity73.8%),”Endurance contraction”of pubococcygeus (PC) was 34.68(AUC:0.60;95%CI,0.51-0.70;sensitivity at 85.1%,specificity33.8%).

Iliococcygeus muscles (IC)

There is no statistical significance in sEMG of Iliococcygeus muscles (IC).

## 4. Discussion

This study is the first attempt to objectively and quantitatively assess the regional PFM function difference between SUI and non-SUI in postmenopausal women through both vaginal digital palpation and the intravaginal physiology-based electrode array airbag probe technology.

Firstly, we used vaginal digital palpation to evaluate pelvic floor muscles according to the modified Oxford grading system, we found the score in SUI in postmenopausal women was lower than non-SUI in postmenopausal women(Table 2). Although it is reported vaginal digital palpation has its limitation, such as it is thought to be subjective and has poor reliability, and lacks the sensitivity to gauge small changes in pressure, while digital palpation should be regularly conducted as one of the methods to evaluate pelvic floor muscles (PFM) according to the International Urogynecological Association and International Continence Society. Our results proved that SUI in postmenopausal women has poor muscle strength compared to non-SUI, both Ignácio Antônio F, et al. [15] and Yang X, et al.[16] have suggested a role for routine clinical digital palpation examination in the evaluation of PFM. According to our study, vaginal digital palpation still a useful and simple strategy for the evaluation of pelvic floor muscles.

In terms of the sEMG acquirement and evaluation technology of PFM, several studies had repoted its value and application in evaluation of PFM in pelvic floor dysfunction [17-19], however, such signals of sEMG is the overall data of the PFM, which can not obtain each specific PFM. We used the novel intravaginal airbag-type electrode array probe to collect the sEMG signals in different PFM regions according to the anatomy position of the PFM, which included urethral sphincter (US), vaginal sphincter (VS), external anal sphincter (EAS), Levator ani muscle (LAM)(Fig 1). Most of the existing vaginal probes can only collect 1-4 channels of sEMG signals with a custom probe[20,21], and their rigid shape cannot adapt multifarious vaginal structure of different women, which makes sEMG-based evaluation not accurate enough. Because of differences in impedance, muscle depth and muscle fiber orientation, Valid comparisons is not possible between groups without normalization. Our method allows the probe inflated to adapt different vaginal structures, and is able to orient the abnormal region location, which cannot be achieved by using the existing technologies. We provide a novel evaluation method for the accurate examination of PFM.

In terms of the specific regional pelvic floor muscle abnormality and/or insufficiency, we found that the significantly different regions between the two groups show that the significantly different muscles between the two groups show obvious difference in muscle strength in sEMG, especially in maximal voluntary contraction(MVC), tonic contractions, endurance contraction, as shown in Table 3 and Figure 2.In the rest station, both in the pre-baseline and post-baseline almost all the PFM showed stable conditions, there is no statistic difference between postmenopausal SUI group and non-SUI group. While, there is a statistically significant decrease in urethral sphincter (US), puborectalis (PR) and pubococcygeus (PC) during flick contractions, tonic contractions and endurance contraction in the postmenopausal SUI group compared with postmenopausal non-SUI group.

The pathogenesis of SUI is associated with anatomical abnormalities involving the urethra, urinary bladder, and urogenital diaphragm. Insufficiency of the urethral sphincter and vesicourethral ligament as well as weakening of the muscle-fascial structures of the whole pelvic floor impairs normal urinary continence.Falah-Hassani K et al [20]have suggested a role for routine clinical EMG examination in the evaluation of urethral sphincter insufficiency. Our ASEA device can provide a tight contact and stable interface between electrode units and PFM and collect high-quality sEMG signals from specific regions of muscles, as shown in Table 3 and Figure2, MVC in US was 43.4 µV vs. 142.88 µV, tonic contractions in US was 24.74µV vs. 47.51µV, Endurance contraction in US was 23.64µV vs. 51.17µV(*p*<0.05), which were lower in the postmenopausal SUI group compared with postmenopausal non-SUI group. Although insufficiency of the urethral sphincter attracted increasing attention, the function of US has always been a difficult problem. Our study provided an access to evaluate the function of US and proved that urethral sphincter insufficiency participated in the pathogenesis of SUI.

Furthermore, Delancey pointed out the “hammock”-like structure formed by the continuity of the intrapelvic fascia, vaginal wall and levator ani muscle. This structure can maintain urethral stability and the urethral closure pressure when the abdominal pressure increases[22]. The Levator ani muscles (LAMs) are considered a functional unit which provides support to the pelvic organs in the transverse plane (lifting) and compresses the urethra against the anterior vagina in the mid-sagittal plane (squeezing). Damage to or dysfunction of the LAMs is thought to be a contributor to SUI. LAM structure and <u>function</u> can be evaluated through many different approaches[20,23,24]. It is accepted that intravaginal dynamometry is recommended as the best approach to directly measure LAM force-generating capacity [20]; yet, as with existing intravaginal device, only combined force contributions are recorded, such measures may not be the precise reflection of pelvic floor muscles characters. Our ASEA device can provide the signals of puborectalis, pubococcygeus, and iliococcygeus muscles separately.

We noted that the sEMG of PFM activity was lower in LAMs, especially in puborectalis (PR) and pubococcygeus (PC) in postmenopausal SUI group. MVC in PR was 45.64 µV vs. 127.19 µV, in PC was 41.76 µV vs. 58.14 µV. Tonic contractions in US was 24.74µV vs. 47.51µV, in PR was 25.75µV vs. 48.51 µV, in PC was 22.36 µV vs. 39.64µV. Lower PFM activity was also noted in US, PR and PC in the postmenopausal SUI group. Endurance contraction in US was 23.64µV vs. 51.17µV, in PR was 23.28µV vs. 52.92 µV, in PC was 23.22 µV vs. 36.86µV. Our study proved that LAMs insufficiency participated in the pathogenesis of SUI, more importantly we proved that puborectalis and pubococcygeus muscles in LAMs which are in the proximity of the center of PFMs, not the whole LAMS played an important role in the pathogenesis of SUI.

In addition, our study firstly used ROC curves to construct separately for all sEMG tests (Figure 3) in order to present the prediction ability of these variables to predict SUI, cutoff point mainly considering results derived from the maximal voluntary contraction (MVC), tonic contractions and endurance contraction tests in US and LAMs.

In urethral sphincter (US), the optimum predict cutoff for MVC was 48.55 (AUC, 0.61; 95% CI, 0.51-0.71; sensitivity at 76.1%, specificity 53.8%, tonic contractions was 25.79 (AUC, 0.61; 95% CI, 0.51-0.70, sensitivity at 77.6%, specificity 44.6%), endurance contraction was 23.02(AUC: 0.61,95%CI:0.53-0.72;sensitivity at 70.1%, specificity 60%). In the article by Ptaszkowski K et al, the optimum diagnostic cutoff for “quick flicks” was 9.15 (AUC, 0.84; 95% CI, 0.78–0.91; sensitivity at 60% specificity, 94%), “contractions” was 11.33 (AUC, 0.80; 95% CI, 0.73–0.87; sensitivity at 60% specificity, 90%), “static hold” was 9.94 [AUC, 0.84;95% CI, 0.78–0.91; sensitivity at 70% specificity, 94%). The sensitivity in each contractions were 60-70%, which is similar to our data, the sensitivity of our data in each contractions in each muscles were as followings: US 70.1%-76.1%, PR 55.2%-74.6%, PC 64.2%-85.1%. while the specificity of this study in each contractions was as high as 90-94%, which is higher than our data. However, such PFM specificity activities were the mean functional PFM activity, all of these methods can not separate specific pelvic muscles to collect signals of each muscles, such as US, PR and PC. These data suggest that measuring sEMG activity in the PFM may be a useful tool to predict the absence of SUI.

## 5. Conclusions

In the population of postmenopausal SUI, we found that the specific functions in US and LAMs, especially in PR and PC were lower than healthy people by our novel airbag-type stretchable electrode array (ASEA) device, which proved that US defect and dysfunctional LAMs play an important role in the pathogenesis of SUI in postmenopausal women. Separation of specific regions of US and LAMs can provide a reliable and optimized treatment strategy for the precise rehabilitation of pelvic floor muscles.

## Data Availability

Deidentified research data will be made publicly available when the study is completed and published.

https://doi.org/10.3390/diagnostics13061158

https://doi.org/10.1002/advs.202004987

## Abbreviations

The following abbreviations are used in this manuscript

ASEA: Novel airbag-type stretchable electrode array
PFM: Pelvic Floor Muscle
sEMG: Surface electromyography
SUI: Stress urinary incontinence
US: Urethral sphincter
LAM: Levator ani muscle
PR: Puborectalis
PC: Pubococcygeus
IC: Iliococcygeus
ARP: Anterior resting potential
MVC: Maximum voluntary contraction
TCP: Tonic contraction potential
ECP: Endurance contraction potential
PRP: Posterior resting potential

## Author Contributions

Conceptualization, Z.J. Z; methodology, Q.C and S.Y. H; software Z.J. Z; validation: Q.C and S.Y. H; formal analysis, Z.J. Z, Q.C and S.Y. H; investigation,W.J. L; data curation, S.M. W; writing—original draft preparation, Z.J. Z; writing—review and editing, Z.J. Z, Q.C and S.Y. H; visualization, Z.J. Z; supervision, Z.W. X; project administration, Z.W. X; funding acquisition, Z.W. X. All authors have read and agreed to the published version of the manuscript.

## Institutional Review Board Statement

The study was conducted in accordance with the Declaration of Helsinki and approved by the Ethics Committee of the Women’s Hospital, School of Medicine, Zhejiang University, Hangzhou, China, (protocol ID: No. IRB 20220008-R (2021)), for studies involving humans.

## Informed Consent Statement

Informed consent was obtained from all subjects involved in the study.

## Data Availability Statement

Data are available upon a request to the authors.

## Acknowledgments

Special thanks to Z.W. X for her guidance through each stage of the process.

